# Regional quantification of cardiac metabolism with hyperpolarized [1 - ^13^ C]-pyruvate MRI evaluated in an oral glucose challenge

**DOI:** 10.1101/2023.10.16.23297052

**Authors:** Peder E. Z. Larson, Shuyu Tang, Xiaoxi Liu, Avantika Sinha, Nicholas Dwork, Sanjay Sivalokanathan, Jing Liu, Robert Bok, Karen G. Ordovas, James Slater, Jeremy W. Gordon, M. Roselle Abraham

**Author notes:** Indicates equal contributions Affiliations (Peder E. Z. Larson, Shuyu Tang). Corresponding author address, 1700 4^th^ Street, Byers Hall Room 102C, San Francisco, CA 94143.

## Abstract

**Background:** The heart has metabolic flexibility, which is influenced by fed/fasting states, and pathologies such as myocardial ischemia and hypertrophic cardiomyopathy (HCM). Hyperpolarized (HP) ^13^C-pyruvate MRI is a promising new tool for non-invasive quantification of myocardial glycolytic and Krebs cycle flux. However, human studies of HP ^13^C-MRI have yet to demonstrate regional quantification of metabolism, which is important in regional ischemia and HCM patients with asymmetric septal/apical hypertrophy.

**Methods:** We developed and applied methods for whole-heart imaging of ^13^C-pyruvate, ^13^C-lactate and ^13^C-bicarbonate, following intravenous administration of [1-^13^C]-pyruvate. The image acquisition used an autonomous scanning method including bolus tracking, real-time magnetic field calibrations and metabolite-specific imaging. For quantification of metabolism, we evaluated ^13^C metabolite images, ratio metrics, and pharmacokinetic modeling to provide measurements of myocardial lactate dehydrogenase (LDH) and pyruvate dehydrogenase (PDH) mediated metabolic conversion in 5 healthy volunteers (fasting & 30 min following oral glucose load).

**Results:** We demonstrate whole heart coverage for dynamic measurement of pyruvate-to-lactate conversion via LDH and pyruvate-to-bicarbonate conversion via PDH at a resolution of 6×6×21 mm^3^ (^13^C-pyruvate) and 12×12×21 mm^3^ (^13^C-lactate, ^13^C-bicarbonate). ^13^C-pyruvate and ^13^C-lactate were detected simultaneously in the RV blood pool, immediately after intravenous injection, reflecting LDH activity in blood. In healthy volunteers, myocardial ^13^C-pyruvate-SNR, ^13^C-lactate-SNR, ^13^C-bicarbonate-SNR, ^13^C-lactate/pyruvate ratio, ^13^C-pyruvate-to-lactate conversion rate, k_PL_, and ^13^C-pyruvate-to-bicarbonate conversion rate, k_PB_, all had statistically significant increases following oral glucose challenge. k_PB_, reflecting PDH activity and pyruvate entering the Krebs Cycle, had the highest correlation with blood glucose levels and was statistically significant.

**Conclusions:** We demonstrate first-in-human regional quantifications of cardiac metabolism by HP ^13^C-pyruvate MRI that aims to reflect LDH and PDH activity.

## Introduction

Cardiac contraction imposes very high energy demands on the heart. The heart is a “metabolic omnivore” able to utilize several substrates (fatty acids, carbohydrates, ketone bodies, amino acids) for ATP generation, but primarily uses fatty acid oxidation (∼40-60%) and glucose oxidation (∼20-40%) at rest. Changes in substrate and oxygen availability, cardiac work, sarcomeric protein gene mutations causing cardiomyopathies^1^ lead to changes in cardiac energy metabolism, which cause secondary changes in gene expression and structural remodeling, that predispose to heart failure and arrhythmias. Hence, in vivo imaging of cardiac metabolism in humans could be helpful for preclinical detection of cardiomyopathy, assessing response to therapies and prognosis.

MRI with hyperpolarized (HP) ^13^C-labeled agents, also known as HP ^13^C MRI, is a new tool for cardiac metabolism imaging^2–6^. It requires no ionizing radiation, and has the unique ability to measure an injected substrate and multiple metabolic products, permitting investigation of myocardial substrate selection and metabolic remodeling. In the case of HP ^13^C-pyruvate MRI, the relative contribution of glucose oxidation and fatty acid oxidation to energy production in the heart can be assessed by measuring pyruvate conversion to [1-^13^C]-bicarbonate, catalyzed by pyruvate dehydrogenase (PDH), and [1-^13^C]-lactate, catalyzed by lactate dehydrogenase (LDH). Initial human cardiac imaging studies of HP ^13^C-pyruvate MRI have demonstrated feasibility^7,8^ and technical improvements^9,10^, as well as the effects of cardiac cycle timing^11^ and adenosine stress tests^12^. Early stage patient imaging studies have shown: decreased myocardial PDH activity measures following chemotherapy in breast cancer patients^13^; reduced PDH and increased LDH activity measures in type 2 diabetes mellitus ^14^; and reduced PDH activity in non-viable tissue post-myocardial infarction^15^. However, these human studies have yet to demonstrate the powerful potential of HP ^13^C MRI to detect and quantify regional differences in cardiac metabolism.

Here, we systematically developed and evaluated methods for regional visualization and quantification of hyperpolarized [1-^13^C] pyruvate MRI. We used a novel autonomous scanning approach for robust and reproducible experiments and, for the first time in humans, demonstrate maps of metabolism across the heart including pharmacokinetic model quantifications that aim to represent PDH and LDH mediated metabolic conversion. We used an oral glucose challenge following fasting to evaluate sensitivity to changes in substrate availability.

## Methods

The key components of the study are illustrated in Figure 1, including the study design, HP ^13^C-pyruvate MRI scanning method, and metabolism quantification methods. All are described in greater detail in the following sections.

**Figure 1:**
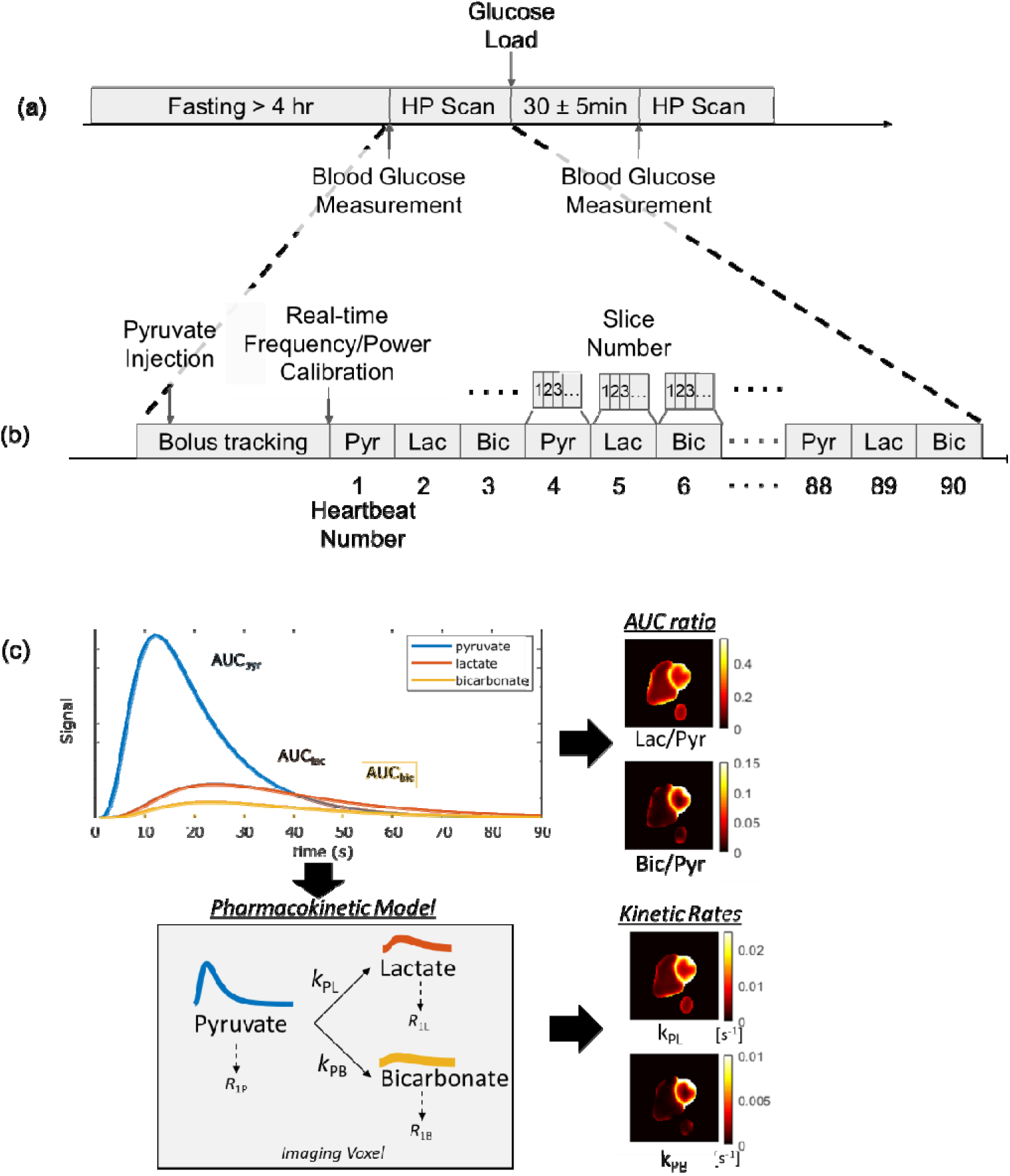
Overview of the (a) study design, (b) HP ^13^C-pyruvate MRI scan, and (c) quantification of metabolism methods.

### Subject recruitment

Seven healthy volunteers with no known cardiovascular disease history or contraindications to MRI were recruited with UCSF Institutional Review Board approval. The subject demographics are shown in Table 1.

**Table 1.**
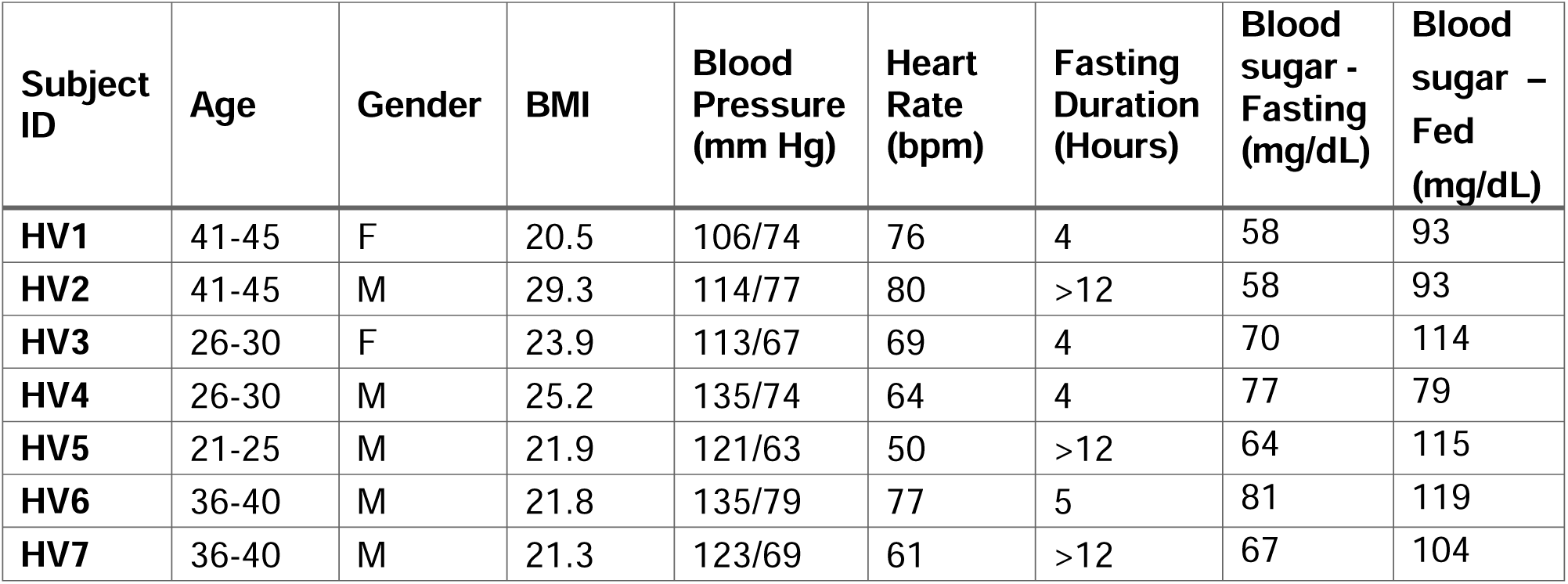
Subject Characteristics at the time of the study including healthy volunteers (HV) who participated in an oral glucose challenge.

| Subject ID | Age | Gender | BMI | Blood Pressure (mm Hg) | Heart Rate (bpm) | Fasting Duration (Hours) | Blood sugar - Fasting (mg/dL) | Blood sugar – Fed (mg/dL) |
| --- | --- | --- | --- | --- | --- | --- | --- | --- |
| HV1 | 41-45 | F | 20.5 | 106/74 | 76 | 4 | 58 | 93 |
| HV2 | 41-45 | M | 29.3 | 114/77 | 80 | >12 | 58 | 93 |
| HV3 | 26-30 | F | 23.9 | 113/67 | 69 | 4 | 70 | 114 |
| HV4 | 26-30 | M | 25.2 | 135/74 | 64 | 4 | 77 | 79 |
| HV5 | 21-25 | M | 21.9 | 121/63 | 50 | >12 | 64 | 115 |
| HV6 | 36-40 | M | 21.8 | 135/79 | 77 | 5 | 81 | 119 |
| HV7 | 36-40 | M | 21.3 | 123/69 | 61 | >12 | 67 | 104 |

### ^13^C pyruvate preparation

For each scan, 1.47g of Good Manufacturing Practices (GMP) [1-^13^C]pyruvate (ISOTEC Stable Isotope Division, MilliporeSigma, Merck KGaA, Miamisburg, OH, USA) mixed with 15mM electron paramagnetic agent (EPA) (AH111501, General Electric Healthcare, Oslo, Norway) was polarized using a 5T SPINlab polarizer (GE, Niskayuna, NY, USA) before being rapidly dissolved with 130°C water and passed through a filter that removed the EPA. Quality-control processes included rapid measurements of pH, pyruvate and EPA concentrations, polarization, and temperature. In parallel, the pyruvate solution was drawn into a syringe (Medrad Inc, Warrendale, PA, USA) through a 0.2 μm sterile filter (ZenPure, Manassas, VA, USA) and transported into the scanner for injection. The delay between dissolution and injection averaged 66.4 s, with a range of 63.4 to 73.1 s. A 0.43 mL/kg dose of an approximately 250 mM hyperpolarized [1-^13^C] pyruvate was injected at a rate of 5mL per second via an intra-venous catheter placed in the antecubital vein, followed by a 20 mL saline flush.

### HP [1-^13^C] pyruvate magnetic resonance imaging

The study design included an oral glucose challenge for the healthy volunteers, and is illustrated in Fig. 1a. Healthy volunteers were asked to fast for at least four hours prior to the study. Two scans with identical intravenous injections of hyperpolarized [1-^13^C] pyruvate and identical imaging protocols were performed on each healthy subject. After the first scan, an oral glucose challenge (drinking a bottle of Gatorade containing 30g total carbohydrates) was performed and followed by a second HP scan starting 30±5 minutes after completing the drink. In all subjects, blood glucose levels were measured with a glucometer (Contour Next, Ascensia Diabetes Care, Parsippany, NJ, USA) less than 5 minutes prior to each hyperpolarized ^13^C injection.

The hyperpolarized MRI scan is illustrated in Fig 1b. It was performed using a volumetric multi-slice, multi-metabolite imaging sequence, acquired dynamically to assess metabolite kinetics following injection. This metabolic imaging was integrated into an autonomous scanning protocol^16^ which included: (1) automatic triggering of the acquisition when the ^13^C-pyruvate bolus arrived in the right ventricle (RV) cavity using low flip angle metabolite-specific imaging; this was followed by (2) a real-time update of the ^13^C frequency using slab-selective MR spectroscopy and (3) measurement and adjustment of the ^13^C transmit power using the Bloch-Siegert B_1_^+^ mapping; and (4) dynamic, multi-slice metabolite-specific imaging (more details below). This autonomous scanning protocol was initiated at the start of the bolus injection and was implemented using the RT-Hawk Research MRI scan control platform (HeartVista, Palo Alto, CA, USA). These automated processes aim to improve the reproducibility of the hyperpolarized ^13^C-MRI scan which must be completed in a rapid (< 2 minute) scan time following bolus injection. Furthermore, frequency and power calibrations are typically performed prior to MRI scans, but for HP ^13^C the natural abundance ^13^C signal available prior to administration is too small to perform these calibrations beforehand. All scanning was performed on a GE 3T MR750 scanner (GE Healthcare, Waukesha, WI, USA).

Short-axis cardiac images of [1-^13^C]pyruvate, [1-^13^C]lactate and ^13^C-bicarbonate were acquired using metabolite-specific imaging sequence that consists of a spectral-spatial excitation and two-dimensional multi-slice cardiac-gated spiral gradient-echo sequence. The ^13^C RF coils consistent of a Helmholz pair “clamshell” transmit coil and an 8-channel “paddle” receive array^17^. Scans used the following parameters: 6 mm x 6 mm in-plane resolution for pyruvate, 12 mm x 12 mm in-plane resolution for bicarbonate and lactate^18^, 21 mm slice thickness, 5 slices, a temporal resolution of 3 heartbeats (∼3.6s), acquired for 90 heartbeats, 20° excitation flip angle for pyruvate, and 30° excitation flip angle for lactate and bicarbonate. The spectral-spatial pulse was 25.17 ms long and excited only a single metabolite at a time. The spiral readout was 22 ms long, and the TR was 57 ms. We simulated expected in-plane resolution based on prior myocardium T_2_* measurements^10^ (also acquired at 3T on GE Healthcare 3T scanner) to be 6.0 mm for pyruvate, 12.5 mm for bicarbonate, and 13.0 mm for lactate (Supplemental Figure S1).

Images were acquired while the subjects were free-breathing. Cardiac triggering was performed automatically by the RT-Hawk software based on the peak signals from a pulse-oximeter placed on a finger. All slices for a single metabolite were acquired starting at a 300 ms delay after each cardiac trigger event, aiming to perform the acquisition during diastole, with an ordering of pyruvate, lactate, bicarbonate repeated 30 times.

### ^1^H MRI

An abbreviated cardiac MR protocol was acquired including localizers to identify the cardiac scan planes and short-axis CINE imaging through the entire heart using a balanced Steady State Free Precession (bSSFP) sequence. CINE image was performed both before and after the HP ^13^C MRI scans. Prior to CINE and the hyperpolarized ^13^C scan, we performed targeted shimming across the ventricles aiming to reduce B_0_ related artifacts for both the ^1^H bSSFP sequences that can suffer from banding artifacts and the ^13^C metabolite-specific imaging that can suffer from reduced flip angles and blurring artifacts^9^.

### Image reconstruction and data analysis

All images were reconstructed using gridding with a Kaiser-Bessel kernel^19,20^ (http://web.stanford.edu/class/ee369c/mfiles/gridkb.m), an oversampling factor of 1.4, and a kernel width of 4.5. Multi-channel data were combined using the method of Roemer^21^ where sensitivity maps were estimated using ^13^C-pyruvate data^22^. Sinc interpolation was used to increase image sizes by a factor of 2 for display by padding the k-space data with zeros prior to transforming; a two-dimensional Fermi window was applied to reduce ringing artifacts. The complex-valued ^13^C images were phased using an assumption of a constant phase offset over the dynamic time series for each voxel. The real component of the resulting phased, complex-valued ^13^C images were used in all subsequent analysis to improve the performance in the low SNR regime, which was particularly important for the bicarbonate measurements.

Quantification of the metabolic images was performed using the dynamic time-series data, Area-under-the-curve (AUC) images, and a pharmacokinetic model, illustrated in Fig 1c. AUC images were calculated by summing the real component of phased images over time. To improve the signal-to-noise ratio (SNR), only a subset of the dynamic time points that contained observable signal in the majority of studies was summed as to not add additional noise. Based on typical dynamics, we chose to compute the pyruvate AUC using the first 25 time-points, lactate AUC using all 30 time-points, and bicarbonate AUC using the 5^th^ to 20^th^ time-points. An AUC SNR is defined as signal magnitude from the AUC maps divided by a noise measurement using a time-point with no signal. This noise measurement was calculated as the standard deviation of the real component from a single time-point after the signal had completely decayed, multiplied by the square root of the number of time-points used in the AUC maps to account for noise averaging in the AUC computation. AUC ratios were computed from the real-valued metabolite AUC maps, avoiding potential bias when using magnitude images that are particularly problematic in the low SNR regime.

Coil-corrected images were created by dividing by measured receive coil sensitivity profiles from an ethylene glycol ^13^C phantom (33 x 20 x 16.5 cm^3^ cylinder) with a proton-density weighted sequence, registered to the in vivo images using fiducial markers placed within the receive coils’ housing^23^. For HP ^13^C, the sparse and inhomogeneous signal distribution in vivo made it challenging to estimate this during the study, motivating the use of prior phantom measurements.

Kinetic rates for conversion from pyruvate to lactate (k_PL_) and pyruvate to bicarbonate (k_PB_) were fit to a uni-directional, three-site pharmacokinetic model with one physical compartment^24^. The real component of the phased ^13^C images were used as input, as using magnitude images creates potential biases especially in the low SNR regime found in our experiments. The model included estimates of signal amplitude and magnetization losses due to the flip angles applied, T_1_ relaxation, and used an “inputless” approach where the measured pyruvate signal is used to calculate the input function^25^. To improve fitting stability, the T_1_ relaxation rates were fixed as T_1,pyruvate_ = 30 s, T_1,lactate_ = 25 s, and T_1,bicarbonate_ = 20 s. There were no constraints on the kinetic rate parameters.

The coil combination, phasing methods, and pharmacokinetic model fitting functions are available in the hyperpolarized-mri-toolbox^26^.

Kinetic rates and AUC ratios were only calculated in voxels that met a minimum pyruvate AUC SNR threshold of approximately 200, which was adjusted for each subject to minimize appearance of artifacts and signal in the lungs.

The LV myocardium was manually segmented on the CINE images at the 300 ms trigger delay to correspond with the HP ^13^C image timing. The LV myocardium segmentation was divided up into a AHA 16-segment model. The corresponding CINE and HP ^13^C images were visually inspected for misregistration, since the CINE data was acquired during breath-holding while the HP ^13^C data was acquired during free-breathing. If necessary, we performed manual rigid registration corrections based aligning the CINE myocardium to the known ^13^C distribution patterns of bicarbonate localized to myocardium and pyruvate having much higher signal in chambers. We did not observe apparent misregistration across the HP ^13^C dynamic time series data.

A paired Students T-test was used to compare ^13^C measurements between fasted and fed states. All measurements used for the T-tests were from real-valued data to avoid the bias from rectification to magnitude images and aiming to preserve the zero-mean Gaussian noise expected in raw MRI data. We confirmed that the metabolite images decayed to zero mean, and also that the AUC and kinetic fits were zero mean in regions without signal. Pearson’s product-moment correlation coefficients, *r*, were computed for the ^13^C measurements as a function of measured blood glucose levels.

## Results

### Human subjects

We studied 7 healthy volunteers ranging in age from 25 to 48 years. Table 1 summarizes subject hemodynamics and blood sugar levels prior to first HP ^13^C MRI scan.

### Cardiac Imaging

Hyperpolarized [1-^13^C] pyruvate MRI was successfully acquired in all 7 subjects. No adverse effects were observed when the subjects were monitored during injection(s) and upon 24-hour follow-up. Images from two healthy volunteers were excluded from the analysis: one due to a technical failure in saving the imaging data, and the other due to low SNR (∼2-3 times lower than all other studies) where we felt we could not reliably quantify ^13^C-bicarbonate. ^1^H MRI imaging was used to delineate LV and RV myocardium, and distinguish myocardium from the cavities (blood pool). We observed good co-registration between anatomic and metabolic imaging. We measured typical ranges of regional wall LV thickness in the healthy volunteers (Supplemental Table S1).

### Metabolite distributions

Figure 2 shows representative hyperpolarized ^13^C AUC images for both the fasting and fed states in a healthy volunteer. ^13^C-pyruvate signal intensity was markedly higher in the RV and LV cavities, when compared to myocardium, and similar in the fed and fasting states. ^13^C-lactate signal intensity was similar in cardiac cavities and myocardium. ^13^C-bicarbonate shows the greatest difference between the fed and fasting states. The bicarbonate signal, reflecting PDH flux, was very low (at the noise level) in the fasting state, and primarily localized to LV myocardium in the fed state. This distribution between the cavities and myocardium was consistent across studies. ^13^C metabolite SNR values are reported in Fig. 6.

**Figure 2.**
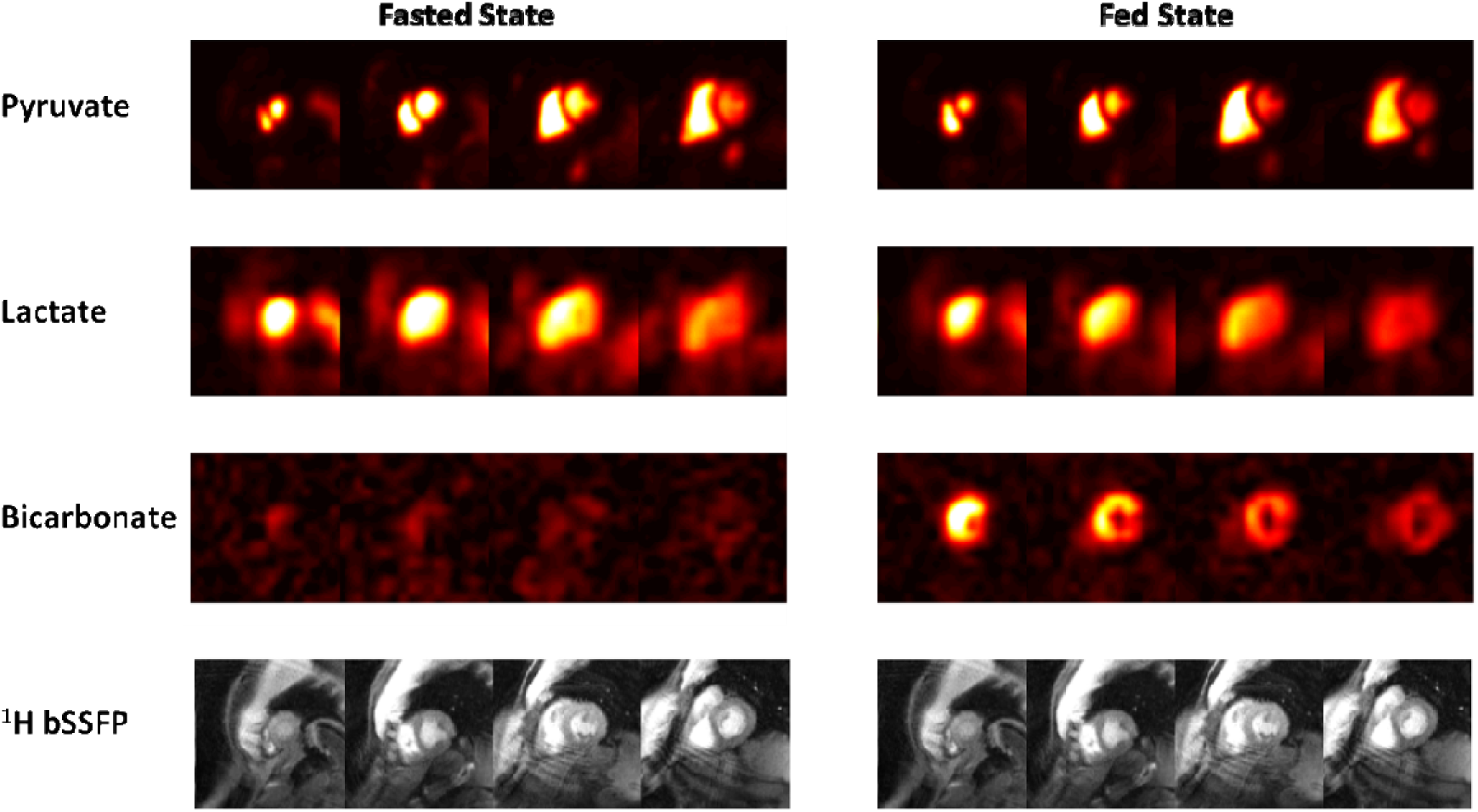
Representative ^13^C AUC images of pyruvate, lactate, and bicarbonate in fasted and fed states with corresponding ^1^H short-axis images in a healthy volunteer (subject HV7). The ^13^C metabolite images (raw) are scaled identically across experiments and slices within each metabolite.

Figure 3 shows representative dynamic ^13^C images of pyruvate, lactate and bicarbonate. As expected for intravenous injection, ^13^C-pyruvate is first observed in RV cavity and then in LV cavity. It appears in the myocardium around 2 s after the bolus arrival, corresponding to tissue perfusion. At early time-points, ^13^C-lactate follows a similar trend, appearing first in the RV cavity, followed by LV cavity, with coincident timing to ^13^C-pyruvate. At the 5^th^ time point, ^13^C-lactate is detectable in the myocardium while still being visible in the blood pool. Since the ^13^C metabolites in the RV cavity must have arrived directly through the veins from the intravenous injection, we hypothesize that the source of this signal is LDH-mediated ^13^C-pyruvate to ^13^C-lactate conversion by red blood cells^27,28^. If this came from incidental excitation of ^13^C-pyruvate by the spectral-spatial pulse during ^13^C-lactate imaging, there would be a spiral off-resonance blurring artifact in the ^13^C-lactate images, but no such artifact is present. ^13^C-bicarbonate is strictly localized to myocardium, appearing at the 5^th^ time point and beyond, similar to ^13^C-lactate dynamics in the myocardium. This indicates that ^13^C-bicarbonate is not originating in blood or outside the heart, and reflects PDH flux in LV myocardium.

**Figure 3.**
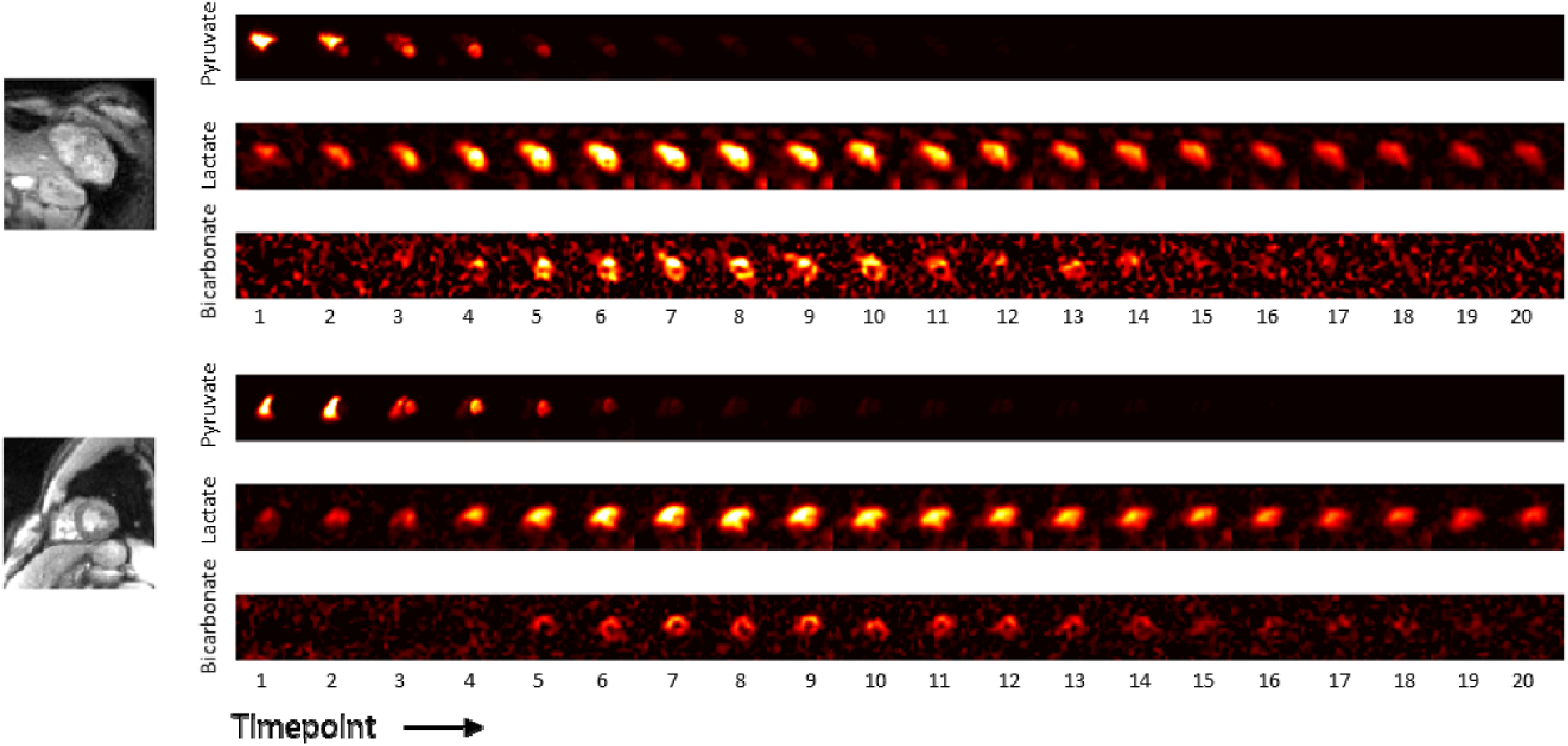
Representative ^13^C multi-slice dynamic images of pyruvate, lactate and bicarbonate in the fed state in two healthy volunteers (subjects HV3, top, and HV6, bottom). Each time-point required 3 heartbeats to acquire, of which the first of each set of 3 heartbeats was used to acquire ^13^C-pyruvate, the second for ^13^C-lactate, and the third for ^13^C-bicarbonate. The ^13^C individual metabolite images are scaled consistently across all time points. ^13^C-pyruvate images are scaled to a third of its maximum for display purposes.

### Quantification of ^13^C data

We investigated 3 strategies to quantify the HP ^13^C images: coil profile correction, ratiometric analysis, and pharmacokinetic modeling. The raw ^13^C images contain substantial signal modulation due to the RF coil sensitivity profiles, particularly from the surface coil receive array used^9^, as can be observed in the previous figures. Following a typical approach used in ^1^H MRI, this effect can be reduced by dividing by an estimate of the coil sensitivity profile, shown in Supplemental Figure S2. However, the resulting homogeneity of the ^13^C images varies across studies. This is likely a result of the difficulty in using sensitivity profiles estimated with a phantom which can be affected by registration, and variations in coil loading and coupling.

Metabolite ratios of ^13^C AUC maps is a common strategy for quantification, as it can correct for RF receive coil profiles since they affect all metabolites the same as well as variations in ^13^C-pyruvate polarization, concentration, and delivery^29^. The ^13^C-lactate/^13^C-pyruvate and ^13^C-bicarbonate/^13^C-pyruvate AUC ratio maps in Fig. 4 show that this approach leads to greater homogeneity of signal distribution in myocardium compared to the metabolite AUC maps in Supplemental Figure S2.

**Figure 4:**
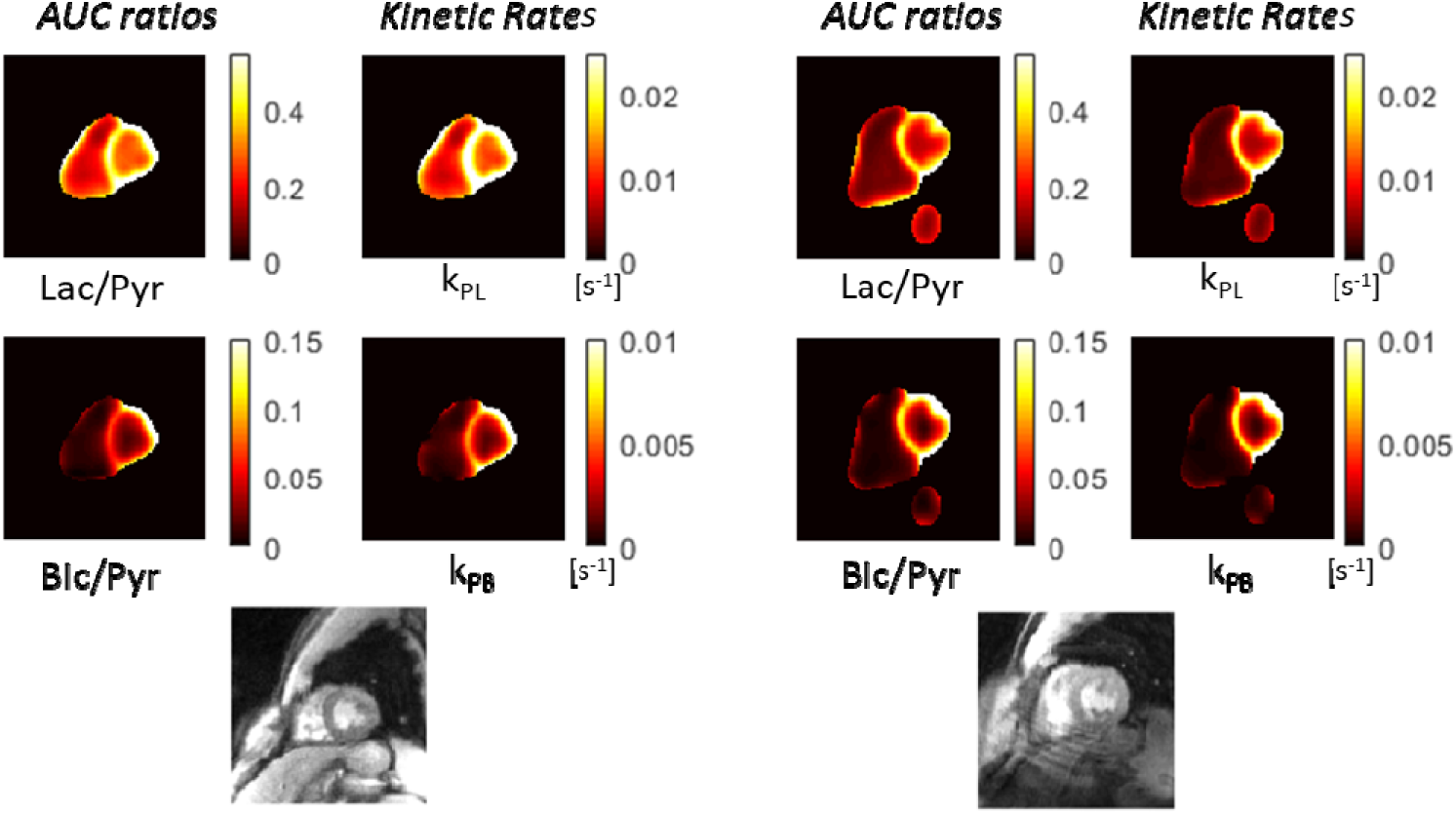
Quantification using AUC ratios and pharmacokinetic modeling in the same healthy volunteer subjects as Fig. 3 (subjects HV6, left, and HV7, right). k_PL_ is the ^13^C-pyruvate to ^13^C-lactate rate and k_PB_ is the ^13^C-pyruvate to ^13^C-bicarbonate rate. Note the maps are only computed in regions that meet a minimum pyruvate AUC SNR threshold as pyruvate signal is required for these quantifications.

Pharmacokinetic modeling is another common strategy for quantification, with the advantage of being more robust across experiments because it can account for the bolus timing, T_1_ relaxation, and flip angle effects. The kinetic rate maps in Fig. 4 show that, similarly to the AUC ratio maps, this leads to a greater homogeneity of signal distribution in myocardium, which we would expect in healthy volunteers.

### Imaging in Fasting and Fed States

Fig. 5 shows the AUC ratios and kinetic rates in all healthy volunteers before and after oral glucose load. In all cases except one, there is uniform increase in the ^13^C-bicarbonate/^13^C-pyruvate and k_PB_ maps in LV myocardium. In subject HV4, metabolite signal is similar in the fed and fasting states, and was associated with little change in blood glucose levels following oral glucose load (see results below).

**Figure 5:**
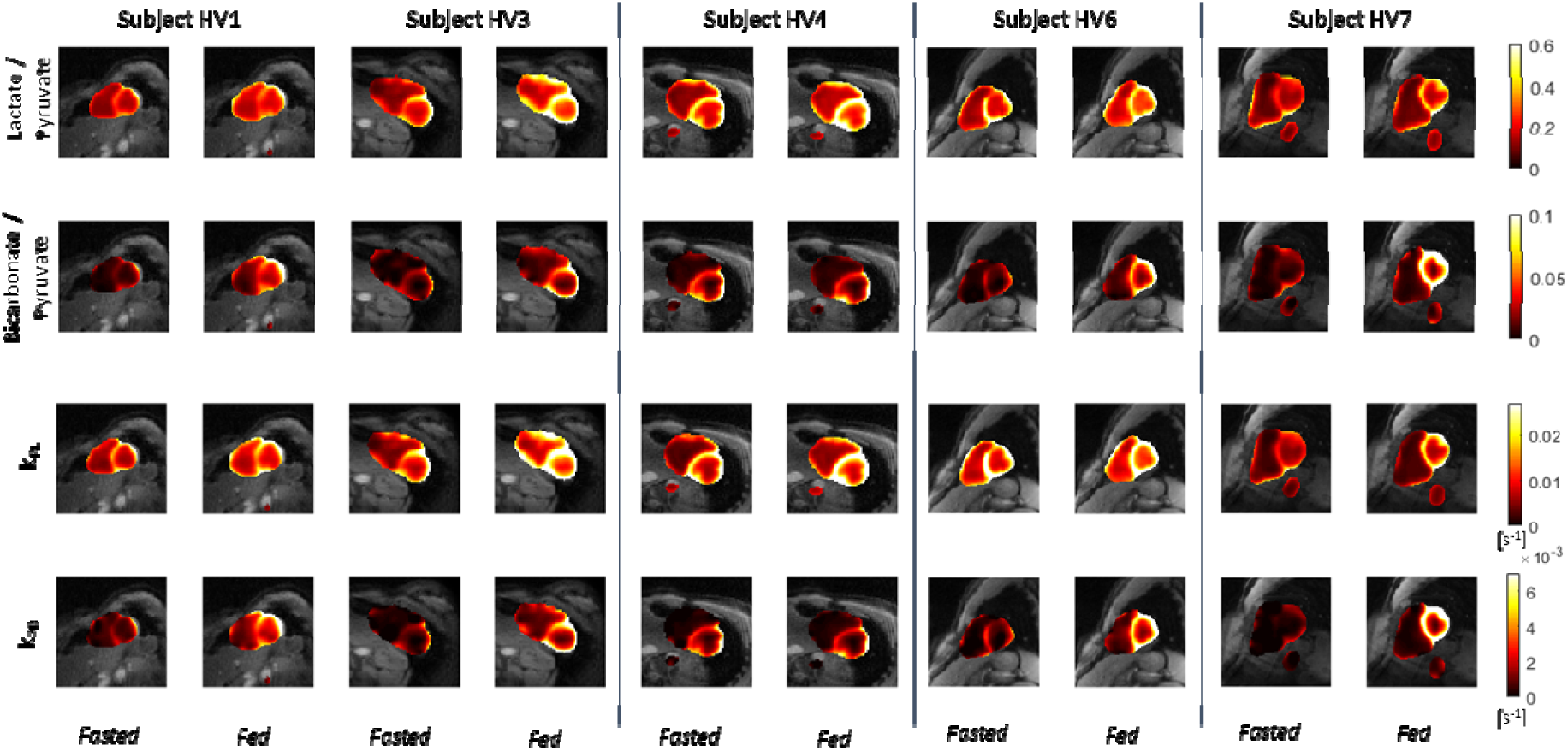
^13^C AUC ratio and kinetic rate maps in healthy volunteers before and after the oral glucose challenge, scaled identically across all scans and overlaid on the ^1^H anatomical images.

Figure 6 shows correlations between blood glucose levels and ^13^C measurements in LV myocardium between fed and fasted states. This data is also shown in Supplemental Tables S2 and S3. There were statistically significant changes in ^13^C-pyruvate SNR, ^13^C-lactate SNR, ^13^C-bicarbonate SNR, ^13^C-lactate/pyruvate, k_PL_ and k_PB_ between the fasted to the fed state (paired T-test, p < 0.05). Notably, subject HV4 who did not have an increase in blood glucose levels following oral glucose challenge also had little change in ^13^C-bicarbonate SNR, ^13^C-bicarbonate/^13^C-pyruvate ratio, and ^13^C-bicarbonate/^13^C-lactate ratio. The strongest correlations with blood glucose levels were observed for ^13^C-bicarbonate SNR (*r* = 0.50), ^13^C-bicarbonate/^13^C-pyruvate ratio (*r* = 0.54) and k_PB_ (*r* = 0.56). The correlation was only statistically significant (p < 0.05) for k_PB_.

**Figure 6:**
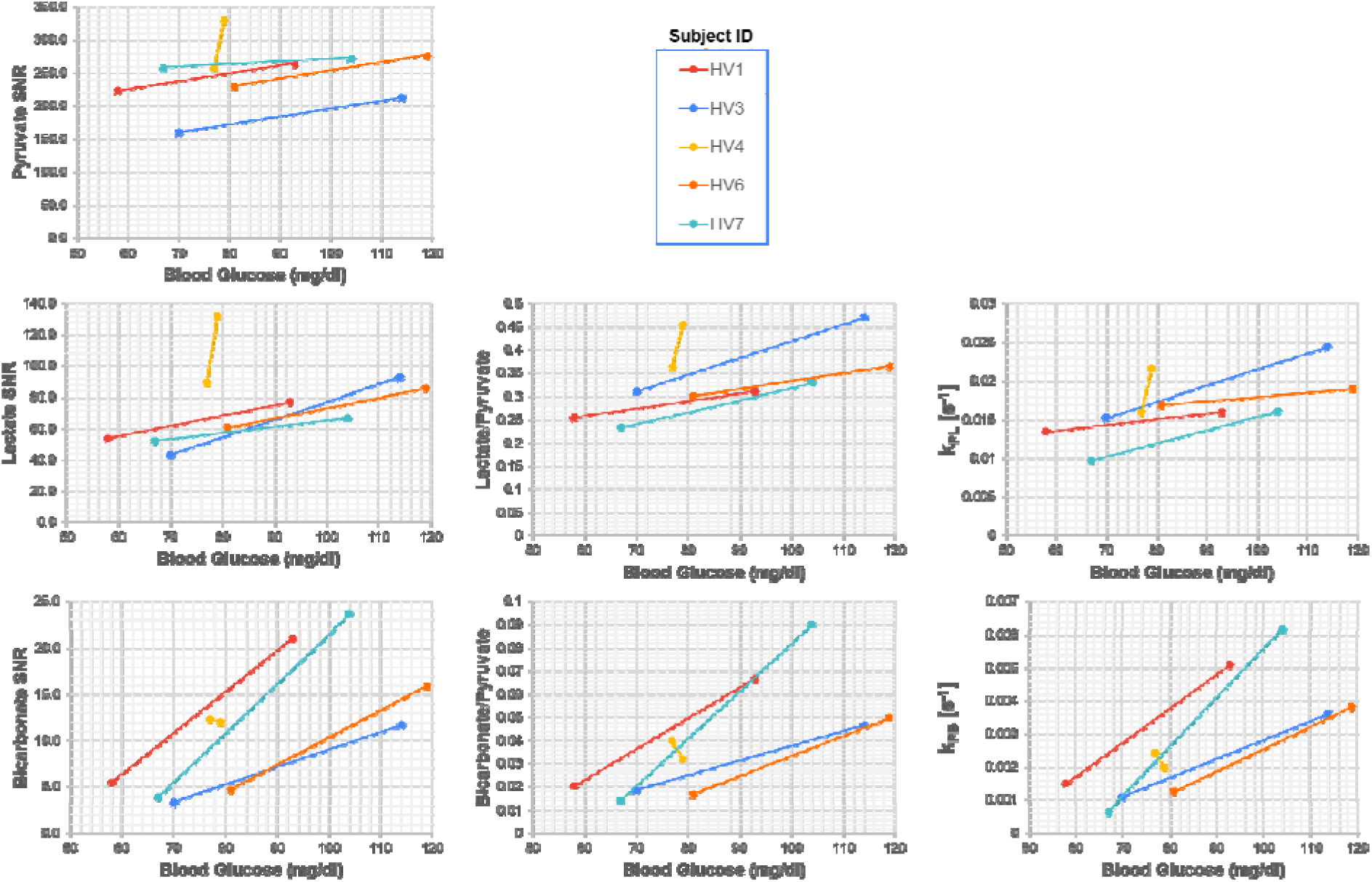
Correlation of hyperpolarized ^13^C measurements in the LV myocardium with blood glucose levels as measured < 5 minutes prior to injection across all healthy volunteer subjects. These data are also shown in Supplementary Tables S2 and S3.

Figure 7 shows how the ^13^C measurements varied spatially, averaged across all studies, using an AHA 16-segment model. The SNR for all metabolites is generally higher in the anterior and septal segments, which we expect based on their proximity to the RF receive coils. The SNR was also higher in the mid and apical segments compared to the basal segments. As in Figure 5, there was strong correspondence between the AUC ratio and kinetic rate metrics. All metrics showed higher average values in the lateral segments, with the highest values in the apical lateral segment. The basal segments had lower values than the mid and apical segments for the metabolism metrics.

**Figure 7:**
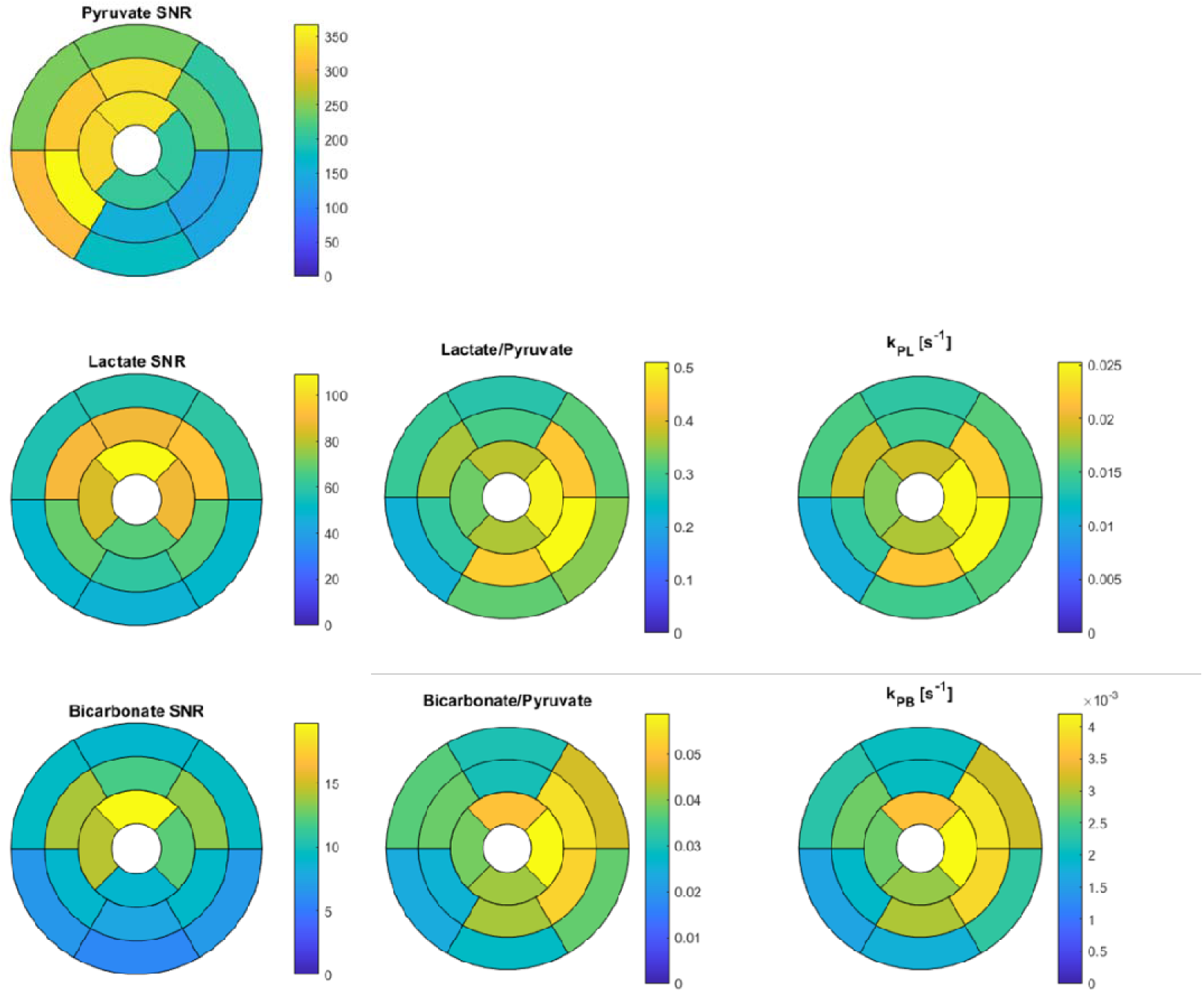
Hyperpolarized ^13^C measurements shown across the myocardium in using an AHA 16-segment model. These data are averages across all subjects and studies shown.

## Discussion

In this study, we demonstrate visualizations and quantifications of LDH and PDH mediated metabolic conversion in the human heart using hyperpolarized ^13^C-pyruvate MRI. The studies consistently achieved coverage of the ventricles for dynamic metabolism measurements that reflect LDH and PDH activity at a resolution of 6x6x21 mm^3^ (pyruvate) and 12×12×21 mm^3^ (lactate, bicarbonate) as well as 16-segment analysis. With these measurements, we are able to spatially resolve metabolism in LV myocardium. There were statistically significant increases in ^13^C-pyruvate SNR, ^13^C-lactate SNR, ^13^C-bicarbonate SNR, ^13^C-lactate/^13^C-pyruvate ratio, ^13^C-bicarbonate/^13^C-pyruvate ratio, and k_PB_ following the oral glucose load in healthy volunteers. Measured blood glucose levels allowed us to control for subject variability (e.g. HV4 did not respond to the oral glucose challenge), and using this data we found that PDH mediated conversion as measured by ^13^C-bicarbonate, ^13^C-bicarbonate/^13^C-pyruvate ratio, and k_PB,_ had the highest correlation with blood glucose levels. The kinetic rate, k_PB_, has the highest correlation coefficient and was the only correlation that was statistically significant. These results demonstrate the ability of HP ^13^C MRI to quantify regional cardiac metabolism, and the initial methods and kinetic rates reported will improve the design and interpretation of future studies.

Previous human heart studies with HP ^13^C MRI have performed crucial demonstrations of feasibility^8^, improvements to pulse sequence and reconstruction methods^9,10^, the effect of cardiac cycle on metabolic imaging^11^, the effect of adenosine stress tests^12^, the effect of fasting and fed states^14^, and patient studies investigating the effects of chemotherapy in breast cancer^13^, type 2 diabetes mellitus^14^, and post-myocardial infarction^15^. Several prior studies have included imaging acquisitions^8–12,15^, ratiometric methods^11,13,14^ and pharmacokinetic modeling^12^. The major unique contribution of this work is to perform regional quantification of metabolism, including comparing ratiometric and pharmacokinetic modeling, with the fed/fasted volunteer studies demonstrating experimental changes in metabolism. Regional quantification of metabolism is important, because obstructive coronary artery disease and HCM can be associated with regional myocardial ischemia and hypertrophy, respectively. Furthermore, it is unknown whether regional hypertrophy commonly seen in HCM is associated with greater degree of metabolic remodeling.

The limitations of this study include a relatively small sample size as well as limited correlative data that maybe necessary to control for confounding factors. Our final analyzed group included 5 subjects and 10 scans, with an age range of 25-44. We also did not capture the diet and exercise capacity of the volunteers, as these are potentially important confounding factors that would affect cardiac metabolism. While our fasting protocol was relatively loose, requiring only a minimum of 4 hours fasting, the purpose of the oral glucose challenge was not to characterize a fully fasted state but simply to modulate substrate availability and observe the effect on metabolic imaging. In fact, we were quite encouraged by the ability of blood glucose to provide reasonable normalization even with this fasting/fed protocol and a non-responder (HV4).

To improve the results in future studies in the human heart, we believe there are several strategies that can be employed based on the current study. Confounding factors of substrate availability as well as individual physiology differences would benefit from additional measurements of substrate levels (e.g. related to fatty acids and ketone bodies) in the blood as well as assessments of diet and exercise capacity. Increased sample sizes, including covering a broader age range as well as diet and exercise habits would also help to investigate potential bias and confounding factors. For subject preparation, most prior studies suggesting imaging in a fed state to allow for PDH measurements by increasing ^13^C-bicarbonate SNR, with animal model studies showing increased conversion to lactate, alanine, and bicarbonate in a fed state^33^. Interestingly, similar performance in terms of sensitivity, repeatability, and measurement of pathologic variations was measured when imaging in fed state created by a carbohydrate rich meal versus an oral glucose load^34^. However, in HCM, imaging in the fasting state may be useful to demonstrate metabolic remodeling in hypertrophied myocardium.

To quantify the HP ^13^C-pyruvate data, both AUC metabolite ratios and kinetic rates showed promise as biomarkers for PDH and LDH flux, revealing higher fluxes in healthy volunteer LV myocardium with patterns that were relatively homogeneous compared to using raw or coil-corrected metabolite images. The pharmacokinetic modeling has theoretical advantages of robust measurements even with different experimental parameters or conditions such as start times, repetition times (proportional to heart rate in this study) and flip angles^25^. With certain conditions, such as capturing of the entire ^13^C signal dynamics or consistent bolus and acquisition timing, the AUC ratios and kinetic rates are proportional^29^. However, in previous HP ^13^C studies of prostate cancer this assumption was shown to be violated primarily due to acquisition and agent delivery timing differences^25^. In this study, the AUC ratio and kinetic rate quantifications were relatively similar, which can likely be attributed to reproducible experimental timings from the bolus triggering method used. We did, however, observe that the kinetic rates had the higher correlation with blood glucose levels than AUC ratios, and k_PB_ was the only parameter where the correlation with blood glucose was statistically significant. We believe this can be attributed to increased robustness of pharmacokinetic modeling to account for residual differences in the start time of the acquisition as well as variations the heart rates. The kinetic rates can also be more readily compared across studies since they account for timing differences as well as protocol changes such as flip angles used.

We observed spatial trends in the segment-based regional analysis of both metabolism quantifications (Fig. 7). For one, the lateral segment metrics were higher on average. One possible explanation could be reduced partial volume effects from pyruvate, where the septal wall in particular could suffer from pyruvate contamination from both the LV and RV blood pools. The apical segments had higher metabolism metrics on average, also where we would expect reduced partial volume effects from pyruvate in the blood. However, we did not observe this increase in anterior segments, which should experience less partial volume effects. These results suggest that the relatively large voxel sizes currently required to achieve adequate SNR for HP ^13^C human heart studies maybe a limiting factor in regional quantification, warranting further improvements in image acquisition, reconstruction, or analyses that incorporate a partial volume effect model.

Interestingly, there was also a statistically significant increase in LV myocardium ^13^C-pyruvate SNR between the fasted and fed state scans. We expect this is not due to experimental factors that influence SNR such as polarization levels, ^13^C-pyruvate dose, flip angles, and RF coil sensitivities, as all of these were either constant or have no expected consistent change from the 1^st^ to 2^nd^ scans. Thus, this increase in ^13^C-pyruvate may reflect increased perfusion and/or uptake of pyruvate in the myocardium, which could be a result with the expected increase in glucose oxidation with higher glucose availability in the fed state.

We had two technical failures in the healthy volunteer studies: in one case (subject HV2), the raw k-space data was not transferred in time from the scanner and was lost, and thus we were unable to perform the optimized coil-combination as well as the image phasing that was crucial for quantifying low metabolite signals (e.g. ^13^C-bicarbonate); in the other case (subject HV5), the SNR was poor (> 2 fold lower than all other studies), which was most likely due to low ^13^C-pyruvate polarization or degraded RF coil performance. In another case (subject HV4), there was no observed difference in HP ^13^C measurements before and after the oral glucose challenge.

We observed clear evidence of ^13^C-lactate in the blood pool at the same time as the ^13^C-pyruvate bolus, with both metabolites first arriving in the RV followed by the LV prior to their appearance in myocardium. Since this is first observed in the RV after intravenous injection, the metabolites had only traveled through the veins from the site of injection to the heart and did not yet circulate anywhere else in the body. This result can be explained by the conversion of ^13^C-pyruvate to ^13^C-lactate via LDH in circulating red blood cells that is occurring as soon as the injection begins^27,28^. This has been shown and debated in several preclinical studies, but to our knowledge, this is the most definitive evidence of this phenomena in humans. This potentially has an impact on all hyperpolarized ^13^C-pyruvate studies, as it implies that the ^13^C-lactate observed can be coming from blood pool metabolic conversion instead of the tissue of interest. This circulating ^13^C-lactate could also be a source of contrast as it can be used as a fuel source^30–32^. It will also introduce partial volume errors in voxels containing myocardium and blood pool, originating from the chambers and potentially coronary arteries, where the additional blood lactate would likely increase the ^13^C-lactate/pyruvate and k_PL_.

## Conclusion

We demonstrated novel regional quantifications of LDH and PDH flux by HP ^13^C MRI in a cohort of healthy volunteers. Both ratiometric and kinetic modeling-based quantifications showed high quality maps with glycolytic metabolism localized to LV myocardium. ^13^C-lactate was also detected in the RV blood pool, immediately after intravenous injection and coincident with ^13^C-pyruvate, reflecting LDH activity in blood, an observation that could significantly affect interpretation of all HP ^13^C MRI studies. In the oral glucose challenge, almost all ^13^C measurements in LV myocardium (pyruvate SNR, lactate SNR, bicarbonate SNR, lactate/pyruvate ratio, k_PL_, and k_PB_) had statistically significant increases following an oral glucose load, with the largest changes in PDH flux measurements. Blood glucose levels were valuable to account for a non-responder to the oral glucose challenge. k_PB_, a measure of PDH flux, had the strongest correlation with blood glucose levels, and was the only measurement with a statistically significant correlation.

## Declarations

### Ethics approval and consent to participate

All participants were recruited with UCSF Institutional Review Board approval (IRB #: 19-27889, PIs: Dr. Maria Roselle Abraham and Dr. Peder Larson) and gave informed consent prior to participation in this study.

### Consent for Publication

All participants data shown gave consent for publication.

### Availability of data and materials

The datasets analyzed during the current study are available from the corresponding author on reasonable request. Much of the data are in included in this published article and its supplementary information files.

## Funding

This work was supported by funding from a UCSF Resource Allocation Program Team Science award, Myokardia Inc. Myoseeds program and NIH grants R33HL161816, P41EB013598, and U24CA253377. ND received post-doctoral training funding from the American Heart Association (grant number 20POST35200152).

### Authors Contributions

PEZL conceived and designed the study, performed data analysis, and interpreted the data. ST created the data acquisition techniques, acquired data, and performed data analysis. XL acquired data and performed data analysis. AS performed data analysis. ND helped create the images. SS performed data analysis. JL helped design the study and performed data analysis. RB helped design the study and oversaw the human subjects recruitment. KGO performed data analysis and interpreted the data. JS oversaw the imaging agent preparation. JWG helped design the data acquisition techniques and assisted with imaging agent preparation. MRA conceived and designed the study, and interpreted the data. All authors read and approved the final manuscript.

## Acknowledgments

We would like to acknowledge: assistance with hyperpolarized experiments from Kimberly Okamoto, Mary Frost, Heather Daniel, James Slater, Andrew Riselli, Evelyn Escobar, and Romelyn Delos Santos; inspiring conversations with Prof. Christoffer Laustsen and Prof. Damien Tyler; and the support of the UCSF Hyperpolarized MRI Technology Resource Center and its leader Prof. Daniel Vigneron.

## References

1. Vakrou S, Abraham MR. Hypertrophic cardiomyopathy: a heart in need of an energy bar? Front Physiol. 2014;5:309. doi:10.3389/fphys.2014.00309

2. Golman K, Petersson JS, Magnusson P, et al. Cardiac metabolism measured noninvasively by hyperpolarized 13C MRI. Magn Reson Med. 2008;59(5):1005–1013. doi:10.1002/mrm.21460

3. Schroeder MA, Lau AZ, Chen AP, et al. Hyperpolarized 13C magnetic resonance reveals early-and late-onset changes to in vivo pyruvate metabolism in the failing heart. European Journal of Heart Failure. 2013;15(2):130–140. doi:10.1093/eurjhf/hfs192

4. Timm KN, Miller JJ, Henry JA, Tyler DJ. Cardiac applications of hyperpolarised magnetic resonance. Progress in Nuclear Magnetic Resonance Spectroscopy. 2018;106–107:66-87. doi:10.1016/j.pnmrs.2018.05.002

5. Wang ZJ, Ohliger MA, Larson PEZ, et al. Hyperpolarized 13C MRI: State of the Art and Future Directions. Radiology. Published online March 5, 2019:182391. doi:10.1148/radiol.2019182391

6. Miller JJ, Lau J, Tyler D. 9 -Hyperpolarized MR in cardiology: probing the heart of life. In: Larson PEZ, ed. Advances in Magnetic Resonance Technology and Applications. Vol 3. Hyperpolarized Carbon-13 Magnetic Resonance Imaging and Spectroscopy. Academic Press; 2021:217-256. doi:10.1016/B978-0-12-822269-0.00006-3

7. Nelson SJ, Kurhanewicz J, Vigneron DB, et al. Metabolic Imaging of Patients with Prostate Cancer Using Hyperpolarized [1-13C]Pyruvate. Sci Transl Med. 2013;5(198):198ra108. doi:10.1126/scitranslmed.3006070

8. Cunningham CH, Lau JYC, Chen AP, et al. Hyperpolarized 13C Metabolic MRI of the Human HeartNovelty and Significance: Initial Experience. Circ Res. 2016;119(11):1177–1182. doi:10.1161/CIRCRESAHA.116.309769

9. Reed GD, Ma J, Park JM, et al. Characterization and compensation of inhomogeneity artifact in spiral hyperpolarized 13C imaging of the human heart. Magnetic Resonance in Medicine. n/a(n/a). 10.1002/mrm.28691

10. Ma J, Chen J, Reed GD, et al. Cardiac measurement of hyperpolarized 13C metabolites using metabolite-selective multi-echo spiral imaging. Magnetic Resonance in Medicine. n/a(n/a). 10.1002/mrm.28796

11. Ma J, Malloy CR, Pena S, et al. Dual-phase imaging of cardiac metabolism using hyperpolarized pyruvate. Magnetic Resonance in Medicine. n/a(n/a). doi:10.1002/mrm.29042

12. Joergensen SH, Hansen ESS, Bøgh N, et al. Detection of increased pyruvate dehydrogenase flux in the human heart during adenosine stress test using hyperpolarized [1-13C]pyruvate cardiovascular magnetic resonance imaging. Journal of Cardiovascular Magnetic Resonance. 2022;24(1):34. doi:10.1186/s12968-022-00860-6

13. Park Jae Mo, Reed Galen D, Liticker Jeff, et al. Effect of Doxorubicin on Myocardial Bicarbonate Production from Pyruvate Dehydrogenase in Women with Breast Cancer. Circulation Research. 0(0). doi:10.1161/CIRCRESAHA.120.317970

14. Rider Oliver J, Apps Andrew, Miller Jack J, et al. Non-Invasive In Vivo Assessment of Cardiac Metabolism in the Healthy and Diabetic Human Heart Using Hyperpolarized 13C MRI. Circulation Research. 0(0). doi:10.1161/CIRCRESAHA.119.316260

15. Apps A, Lau JYC, Miller JJJJ, et al. Proof-of-Principle Demonstration of Direct Metabolic Imaging Following Myocardial Infarction Using Hyperpolarized 13C CMR. JACC Cardiovasc Imaging. 2021;14(6):1285–1288. doi:10.1016/j.jcmg.2020.12.023

16. Tang S, Milshteyn E, Reed G, et al. A regional bolus tracking and real-time B1 calibration method for hyperpolarized 13 C MRI. Magn Reson Med. 2019;81(2):839–851. doi:10.1002/mrm.27391

17. Tropp J, Lupo JM, Chen AP, et al. Multi-Channel Metabolic Imaging, with SENSE reconstruction, of Hyperpolarized [1-13C] Pyruvate in a Live Rat at 3.0 tesla on a Clinical MR Scanner. J Magn Reson. 2011;208(1):171–177. doi:10.1016/j.jmr.2010.10.007

18. Gordon JW, Autry AW, Tang S, et al. A variable resolution approach for improved acquisition of hyperpolarized 13 C metabolic MRI. Magn Reson Med. 2020;84(6):2943–2952. doi:10.1002/mrm.28421

19. P. J. Beatty, D. G. Nishimura, J. M. Pauly. Rapid gridding reconstruction with a minimal oversampling ratio. IEEE Trans\ Med\ Imaging. 2005;24(6):799–808.

20. Jackson JI, Meyer CH, Nishimura DG, Macovski A. Selection of a Convolution Function for Fourier Inversion Using Gridding. IEEE Trans\ Med\ Imaging. 1991;10(3):473–478.

21. Roemer PB, Edelstein WA, Hayes CE, Souza SP, Mueller OM. The NMR phased array. Magnetic Resonance in Medicine. 1990;16(2):192–225. doi:10.1002/mrm.1910160203

22. Zhu Z, Zhu X, Ohliger MA, et al. Coil combination methods for multi-channel hyperpolarized 13C imaging data from human studies. Journal of Magnetic Resonance. 2019;301:73–79. doi:10.1016/j.jmr.2019.01.015

23. Dominguez-Viqueira W, Geraghty BJ, Lau JYC, Robb FJ, Chen AP, Cunningham CH. Intensity correction for multichannel hyperpolarized 13C imaging of the heart. Magn Reson Med. 2016;75(2):859–865. doi:10.1002/mrm.26042

24. Zierhut ML, Yen YF, Chen AP, et al. Kinetic modeling of hyperpolarized 13C1-pyruvate metabolism in normal rats and TRAMP mice. J Magn Reson. 2010;202(1):85–92. doi:10.1016/j.jmr.2009.10.003

25. Larson PEZ, Chen HY, Gordon JW, et al. Investigation of analysis methods for hyperpolarized 13C-pyruvate metabolic MRI in prostate cancer patients. NMR Biomed. 2018;31(11):e3997. doi:10.1002/nbm.3997

26. Hyperpolarized-MRI-Toolbox. doi:10.5281/zenodo.1198915

27. Brindle KM, Campbell ID, Simpson RJ. A 1H-NMR study of the activity expressed by lactate dehydrogenase in the human erythrocyte. Eur J Biochem. 1986;158(2):299–305.

28. Romijn JA, Chinkes DL, Schwarz JM, Wolfe RR. Lactate-pyruvate interconversion in blood: implications for in vivo tracer studies. Am J Physiol. 1994;266(3 Pt 1):E334-40.

29. Hill DK, Orton MR, Mariotti E, et al. Model free approach to kinetic analysis of real-time hyperpolarized 13C magnetic resonance spectroscopy data. PLoS One. 2013;8(9):e71996. doi:10.1371/journal.pone.0071996

30. Lau AZ, Chen AP, Cunningham CH. Cardiac metabolic imaging using hyperpolarized [1-13C]lactate as a substrate. bioRxiv. Published online December 6, 2020:2020.12.04.412296. doi:10.1101/2020.12.04.412296

31. Mayer D, Yen YF, Josan S, et al. Application of hyperpolarized [1-^13^C]lactate for the in vivo investigation of cardiac metabolism. NMR Biomed. 2012;25(10):1119–1124. doi:10.1002/nbm.2778

32. Chen AP, Lau JYC, Alvares RDA, Cunningham CH. Using [1-(13) C]lactic acid for hyperpolarized (13) C MR cardiac studies. Magn Reson Med. 2015;73(6):2087–2093. doi:10.1002/mrm.25354

33. Tougaard RS, Szocska Hansen ES, Laustsen C, et al. Hyperpolarized [1-13 C]pyruvate MRI can image the metabolic shift in cardiac metabolism between the fasted and fed state in a porcine model. Magn Reson Med. 2019;81(4):2655–2665. doi:10.1002/mrm.27560

34. Timm KN, Apps A, Miller JJ, et al. Assessing the optimal preparation strategy to minimize the variability of cardiac pyruvate dehydrogenase flux measurements with hyperpolarized MRS. NMR in Biomedicine. 2018;31(9):e3992. doi:10.1002/nbm.3992

