## Supplemental Information for "Regional quantification of cardiac metabolism with hyperpolarized [1 - ^13^ C]-pyruvate MRI evaluated in an oral glucose challenge"

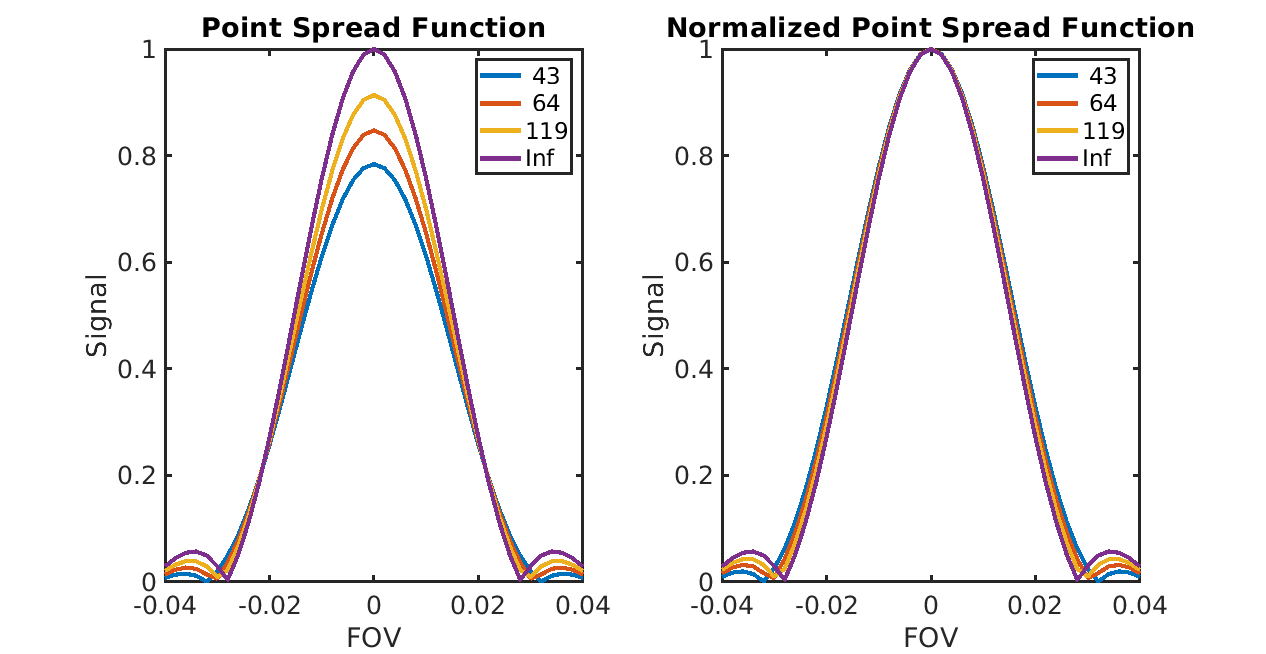
**Supplemental Figure S1:** Point spread functions for the 22 ms long single-shot spiral readout trajectory, using the average T2* values reported from Ma J, Chen J, Reed GD, et al. Cardiac measurement of hyperpolarized 13C metabolites using metabolite-selective multi-echo spiral imaging. Magnetic Resonance in Medicine. https://doi.org/10.1002/mrm.28796 (values noted in the figure) of T2* = 119 ms (pyruvate), T2* = 43 ms (lactate), and T2* = 64 ms (bicarbonate), compared to the ideal point spread function (T2* = “Inf”). These values were measured at the same field strength (3T) and from the same scanner vendor (GE Healthcare) as our study. Based on this, we estimated relative full-width half-maximum (FWHM) for pyruvate = 1.02, lactate = 1.06, and bicarbonate = 1.04, corresponding to effective resolutions of 6.1 mm for pyruvate, 12.7 mm for lactate, and 12.5 mm for bicarbonate in our studies.

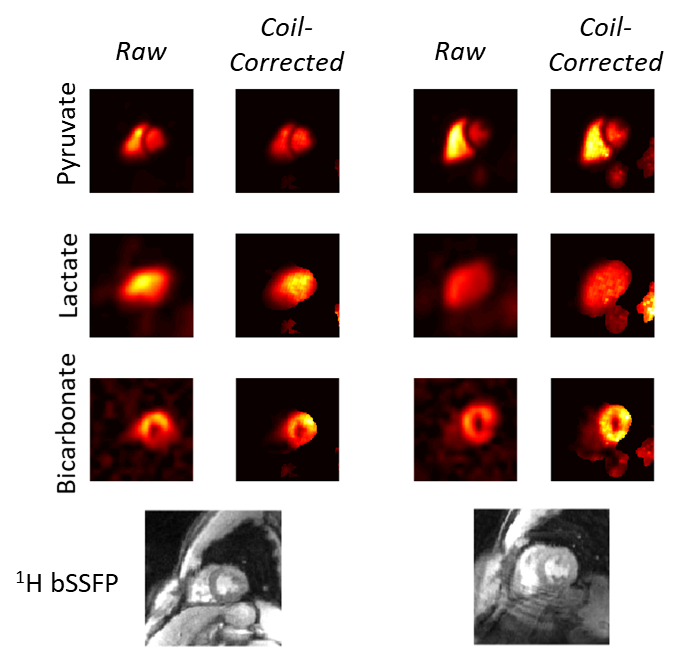

**Supplemental Figure S2:** Examples of coil-corrected HP ^13^C AUC metabolite maps in healthy volunteer subjects (subjects HV6, left, and HV7, right). Note the coil-corrected maps are masked based on a signal threshold to reduce visualization of noise amplification.

| **Subject ID** | **Basal Septum**  **[cm]** | **Mid Septum**  **[cm]** | **Basal Anterior [cm]** | **Mid**  **Anterior**  **[cm]** | **Basal**  **Lateral**  **[cm]** | **Mid**  **Lateral**  **[cm]** | **Apex**  **[cm]** | **Inferior**  **[cm]** |
| --- | --- | --- | --- | --- | --- | --- | --- | --- |
| **HV1** | 8.3 | 5.1 | 5.2 | 4.6 | 5.0 | 4.9 | 3.7 | 3.7 |
| **HV3** | 6.8 | 6.2 | 5.8 | 4.6 | 6.3 | 5.7 | 5.2 | 4.9 |
| **HV4** | 7.6 | 6.2 | 5.5 | 4.3 | 6.2 | 6.1 | 4.5 | 4.1 |
| **HV6** | 8.0 | 7.6 | 6.3 | 4.6 | 6.2 | 6.2 | 6.2 | 3.6 |
| **HV7** | 7.6 | 8.0 | 6.0 | 5.6 | 7.1 | 5.8 | 5.2 | 3.9 |

**Supplemental Table S1**: Regional wall LV thickness, measured from Cine MRI, in all subjects that were included in the analysis.

| **Subject ID** | **ROI** | **^13^C-bicarbonate /^13^C-pyruvate ratio** | | **k_PB_ [s^-1^]** | |
| --- | --- | --- | --- | --- | --- |
|  |  | Fasting | Fed | Fasting | Fed |
| **HV1** | Septum | 0.014 | 0.042 | 0.0010 | 0.0032 |
|  | Global LV | 0.020 | 0.067 | 0.0015 | 0.0051 |
| **HV3** | Septum | 0.013 | 0.033 | 0.0007 | 0.0026 |
|  | Global LV | 0.018 | 0.047 | 0.0011 | 0.0036 |
| **HV4** | Septum | 0.031 | 0.026 | 0.0019 | 0.0015 |
|  | Global LV | 0.040 | 0.032 | 0.0024 | 0.0020 |
| **HV6** | Septum | 0.016 | 0.038 | 0.0014 | 0.0028 |
|  | Global LV | 0.017 | 0.050 | 0.0012 | 0.0038 |
| **HV7** | Septum | 0.009 | 0.061 | 0.0004 | 0.0039 |
|  | Global LV | 0.014 | 0.090 | 0.0006 | 0.0061 |

**Supplemental Table S2**: Quantifications of PDH flux from HP ^13^C-pyruvate MRI, measured across several ROIs in all subjects that were included in the analysis.

| **Subject ID** | **ROI** | **^13^C-lactate /^13^C-pyruvate ratio** | | **k_PL_ [s^-1^]** | |
| --- | --- | --- | --- | --- | --- |
|  |  | Fasting | Fed | Fasting | Fed |
| **HV1** | Septum | 0.213 | 0.252 | 0.0115 | 0.0128 |
|  | Global LV | 0.255 | 0.311 | 0.0135 | 0.0160 |
| **HV3** | Septum | 0.238 | 0.412 | 0.0120 | 0.0214 |
|  | Global LV | 0.310 | 0.470 | 0.0153 | 0.0243 |
| **HV4** | Septum | 0.347 | 0.416 | 0.0148 | 0.0197 |
|  | Global LV | 0.363 | 0.454 | 0.0160 | 0.0215 |
| **HV6** | Septum | 0.303 | 0.338 | 0.0189 | 0.0184 |
|  | Global LV | 0.300 | 0.365 | 0.0169 | 0.0189 |
| **HV7** | Septum | 0.208 | 0.256 | 0.0086 | 0.0117 |
|  | Global LV | 0.234 | 0.328 | 0.0097 | 0.0161 |

**Supplemental Table S3**: Quantifications of LDH flux from HP ^13^C-pyruvate MRI, measured across several ROIs in all subjects that were included in the analysis.
